# Assessment of Potential Adverse Events Following the 2022–2023 Seasonal Influenza Vaccines Among U.S. Adults Aged 65 Years and Older

**DOI:** 10.1101/2023.09.20.23295817

**Authors:** Xiangyu (Chianti) Shi, Joann F. Gruber, Michelle Ondari, Patricia C. Lloyd, Pablo Freyria Duenas, Tainya C. Clarke, Gita Nadimpalli, Sylvia Cho, Laurie Feinberg, Mao Hu, Yoganand Chillarige, Jeffrey A. Kelman, Richard A. Forshee, Steven A. Anderson, Azadeh Shoaibi

## Abstract

**Background:** While safety of influenza vaccines is well-established, some studies have suggested potential associations between influenza vaccines and certain adverse events (AEs). This study examined the safety of the 2022–2023 influenza vaccines among U.S. adults **≥** 65 years.

**Methods:** A self-controlled case series compared incidence rates of anaphylaxis, encephalitis/encephalomyelitis, Guillain Barré-Syndrome (GBS), and transverse myelitis following 2022–2023 seasonal influenza vaccinations (i.e., any, high-dose or adjuvanted) in risk and control intervals among Medicare beneficiaries **≥** 65 years. We used conditional Poisson regression to estimate incidence rate ratios (IRRs) and 95% confidence intervals (CIs) adjusted for event-dependent observation time, seasonality, and outcome misclassification. For AEs with any statistically significant associations, we stratified results by concomitant vaccination status.

**Results:** Among 12.7 million vaccine recipients, we observed 76 anaphylaxis, 276 encephalitis/encephalomyelitis, 134 GBS and 75 transverse myelitis cases. Only rates of anaphylaxis were elevated in risk compared to control intervals. With all adjustments, an elevated, but non-statistically significant, anaphylaxis rate was observed following any (IRR: 2.40, 95% CI: 0.96–6.03), high-dose (IRR: 2.31, 95% CI: 0.67–7.91), and adjuvanted (IRR: 3.28, 95% CI: 0.71–15.08) influenza vaccination; anaphylaxis IRRs were 2.54 (95% CI: 0.49–13.05) and 1.64 (95% CI: 0.38–7.05) for those with and without concomitant vaccination, respectively.

**Conclusions:** Rates of encephalitis/encephalomyelitis, GBS, or transverse myelitis were not elevated following 2022–2023 seasonal influenza vaccinations among U.S. adults **≥** 65 years. There was an increased rate of anaphylaxis following influenza vaccination that may have been influenced by concomitant vaccination.

## 1. Background

Influenza vaccines are the primary tool used to prevent influenza-related hospitalizations and deaths. These vaccines are particularly important for persons aged 65 years and older who are disproportionately affected by severe influenza.^(1, 2)^ Several influenza vaccines were approved for the 2022–2023 influenza season, with three preferentially recommended by the Centers for Disease Control and Prevention for people aged 65 years and older: Fluzone High-Dose Quadrivalent inactivated, Flublok Quadrivalent recombinant, and Fluad Quadrivalent adjuvanted vaccines.^(1)^ As of February 2023, vaccination coverage for the 2022–2023 influenza season among Medicare Fee-For-Service beneficiaries 65 years and older was 53.7% representing the highest influenza vaccination rate among individuals above 18 years of age.^(3, 4)^

Influenza vaccines have been well-established as safe; multiple studies over various influenza seasons have found no association between several adverse events (AEs) and influenza vaccines.^(5-9)^ Certain case reports and epidemiological studies have, however, suggested an association between certain seasonal influenza vaccines and AEs including anaphylaxis, encephalitis/encephalomyelitis, Guillain Barré-Syndrome (GBS) and transverse myelitis.^(10-16)^ Each year influenza vaccines are newly formulated to match several of the expected influenza strains in the United States (U.S.) for the upcoming season. Accordingly, monitoring of rare AEs is important to ensure safety, particularly among persons 65 years and older who may be more vulnerable to AEs because of comorbidities and declining immune function.^(2)^

The U.S. Food and Drug Administration (FDA) Center for Biologics Evaluation and Research monitors the safety of vaccines, including influenza vaccines. As part of the collaboration between FDA and Center for Medicare & Medicaid Services (CMS), we assessed the safety of the 2022–2023 influenza vaccines among Medicare beneficiaries 65 years and older using a self-controlled case series design. The primary objective evaluated the association between influenza vaccination and anaphylaxis, encephalitis/encephalomyelitis, GBS, and transverse myelitis. The secondary objective examined the effect of concomitant vaccination for AEs with any statistically significant association in primary analyses.

## 2. Methods

### 2.1 Study Population, Data Source, and Study Period

Our study included Medicare Fee-for-Service beneficiaries 65 years and older who received an age-appropriate influenza vaccine approved for the 2022–2023 influenza season. To reduce data lag, we used the Medicare Shared Systems Data capturing administrative claims data prior to final adjudication. The Medicare enrollment database captured demographic characteristics. Medicare Fee-For-Service and Part D claims were used to identify individuals’ vaccination history, health characteristics and service utilization. Finally, the Minimum Data Set 3.0 was used to determine nursing home residency status.

The study period was August 1, 2022–June 2, 2023. The study period was defined to align with the start of the influenza season and to ensure adequate vaccine capture and 90% data completeness of AEs.

### 2.2 Study Design and Follow-Up Time

We used a self-controlled case series design to compare AE incidence during risk and control intervals. Time-invariant confounders were controlled for using this design, because comparisons were made within, rather than between, individuals.^(17)^ The risk interval varied by AE (Table 1) and was the period of hypothesized excess risk following vaccination. The control interval was defined as all other follow-up time in the 90-day observation period following vaccination that was not in the risk interval, with the exception of anaphylaxis.^(17)^ For anaphylaxis, the observation period was 16 days starting from vaccination date; the second day following vaccination was excluded from both risk and control intervals, because based on evidence, it is unclear if anaphylaxis risk related to vaccination could persist into this day.^(18, 19)^ Observation periods were censored at the earlier of disenrollment, death, subsequent influenza vaccination, or study period end. Only study participants contributing time to both risk and control intervals were included in the self-controlled case series analysis.

**Table 1.** Adverse event definitions and specifications.

| Adverse Event | Adverse Event Specifications |  |  |  |  |
| --- | --- | --- | --- | --- | --- |
|  | ICD-10 CM codes | Care Setting <sup>a</sup> | Risk Interval (days) | Control Interval (days) | Clean Interval (days) <sup>b</sup> |
| Anaphylaxis | T782XXA,<br>T8052XA | IP, OP-ED | 0-1 | 3-16 | 30 |
| Encephalitis/<br>Encephalomyelitis | G0400, G0402,<br>G0481, G0490,<br>G053 | IP | 1-42 | 43-90 | 183 |
| Guillain-Barré Syndrome | G610 | IP (primary diagnosis<br>position) | 1-42 | 43-90 | 365 |
| Transverse Myelitis | G373 | IP, OP-ED | 1-42 | 43-90 | 365 |

Acronyms: IP – Inpatient; OP-ED – Outpatient Emergency Department.
<sup>a</sup> Care setting definitions: IP refers to inpatient facility claims. OP-ED refers to a subset of outpatient facility claims occurring in the emergency department. OP/PB refers to all outpatient facility claims, and professional/provider claims except those with a laboratory place of service.
<sup>b</sup> Duration prior to the first adverse event (AE) occurrence in the observation period, persons with an observed AE during this period were excluded from the study population. This ensured only incident cases of the event were captured. Cleaning occurred relative to the incident AE occurrence date, and within AE-specific care settings.

### 2.3 Exposures and Adverse Events

The study included influenza vaccines indicated for persons aged 65 years and older in the U.S. during the 2022–2023 influenza season. Vaccine administrations were identified using Current Procedural Terminology (CPT) codes authorized for billing 2022–2023 influenza vaccines.^(20)^ Only first influenza vaccinations during the study period and prior to November 12, 2022 (vaccine cutoff date) were included. The vaccine cutoff date ensured sufficient capture of influenza vaccines and adequate follow-up time to observe events during the full duration of risk and control intervals with at least 90% data completeness. We excluded persons with multiple influenza vaccinations within three days of each other, because we could not determine their exact vaccination date.

Anaphylaxis, encephalitis/encephalomyelitis, GBS, and transverse myelitis were selected as AEs (Table 1) based on their potential association with influenza vaccines determined through literature review and clinical consultation.^(10-16)^ Each outcome was defined based on a claims-based algorithm. To include only incident AEs post-vaccination, persons entered AE-specific cohorts only if the AE did not occur during the clean interval prior to the post-vaccination AE occurrence.

### 2.4 Statistical Analyses

We summarized case frequency stratified by demographic and clinical characteristics. For primary analyses, we estimated incidence rate ratios (IRRs) and 95% confidence intervals comparing AE rates in the risk compared to control intervals within the same individual using conditional Poisson regression. For each AE, the attributable risk (AR) per 100,000 vaccine doses was also estimated to assess the number of excess cases predicted from the regression model divided by the number of eligible vaccinations. Primary analyses were conducted for those with any age-appropriate influenza vaccine and also stratified by high-dose and adjuvanted vaccine groups. Secondary analyses were performed for AEs with statistically significant associations from primary analyses, to evaluate the potential influence of same-day concomitant vaccination. Rates of AEs were separately estimated among individuals that received any of the prespecified concomitant vaccines on the influenza vaccination date, and those who did not receive any concomitant vaccine. Concomitant vaccines of interest included the bivalent COVID-19 mRNA (BNT162b2 bivalent and mRNA-1273.222), hepatitis B, pneumococcal conjugate (PCV15/20, PPSV23), tetanus, and zoster vaccines.^(20)^ These vaccines were selected based on their potential association with the study AEs and their indication in persons 65 years and older.^(21-24)^ Secondary analyses were not stratified by high-dose or adjuvanted vaccine groups because of limited sample sizes.

Both primary and secondary analyses included adjustments to account for potential bias including event-dependent observation time, seasonality, and outcome misclassification (where feasible). The adjustment for event-dependent observation time developed by Farrington et al. was used to adjust for curtailed observation time due to AE-related case fatality.^(25)^ The highest case fatality rate for any outcome was for encephalitis/encephalomyelitis, which had a 14% case fatality rate. In addition, we adjusted for seasonality to reduce bias from seasonal AE patterns.^(26, 27)^ The seasonality adjustment used the daily incidence rates of adverse events estimated from corresponding calendar months in 2021 among the Medicare population 65 years and older, as the baseline incidence for risk and control intervals. To adjust for outcome misclassification from our claims-based AE definitions, a positive predictive value (PPV)-adjusted analysis was performed. The PPV was defined as the proportion of true cases among cases adjudicated through prior medical record review (MRR) following COVID-19 vaccination. PPV estimates were available for anaphylaxis (PPV: 66%, 95% CI: 56–76%)^(28)^ and GBS (PPV: 71%, 95% CI: 63–79%).^(29)^ No MRR was conducted for encephalitis/encephalomyelitis and transverse myelitis at the time of the study, thus PPV estimates were unavailable. For anaphylaxis and GBS, multiple imputation was used to adjust for outcome misclassification by creating multiple datasets where the chart-confirmed status of cases was imputed using the PPV.^(30)^ Statistical analyses were performed on these imputed datasets, and IRR and AR measures combined. Seasonality and PPV adjustments were implemented separately and concurrently where possible.

A post-hoc patient claims profile analysis was performed by two clinicians to investigate the performance of the claims-based transverse myelitis algorithm in detecting incident idiopathic transverse myelitis cases (i.e., cases possibly attributable to vaccination rather than potential unrelated causes). Supplementary Table 1 details the approach further.

All analyses were conducted using R 4.0.3 (R Foundation for Statistical Computing, Vienna, Austria), and SAS v. 9.4 (SAS Institute Inc., Cary, NC, United States).

This surveillance activity was conducted as part of the FDA public health surveillance mandate.

## 3. Results

### 3.1 Descriptive Results

A total of 12.7 million influenza vaccine recipients and doses were captured in the study with each person contributing a single dose (Supplementary Table 2). Among eligible vaccine recipients, we observed 76 anaphylaxis, 276 encephalitis/encephalomyelitis, 134 GBS, and 75 transverse myelitis cases (Supplementary Table 2).

Table 2 presents case characteristics for each of the AE study cohorts. Our case populations consisted of similar proportions of males (46–51%) and females (49–54%), and most persons were white (84–88%). There was a high prevalence of immunocompromised persons (45–76%). No influenza diagnoses were observed in the 30 days prior to vaccination among cases. The prevalence of concomitant vaccination on the same day as influenza vaccination ranged from 25–34% with the majority of coadministrations occurring with bivalent COVID-19 mRNA vaccines (74–92% of concomitant vaccinations).

**Table 2.**
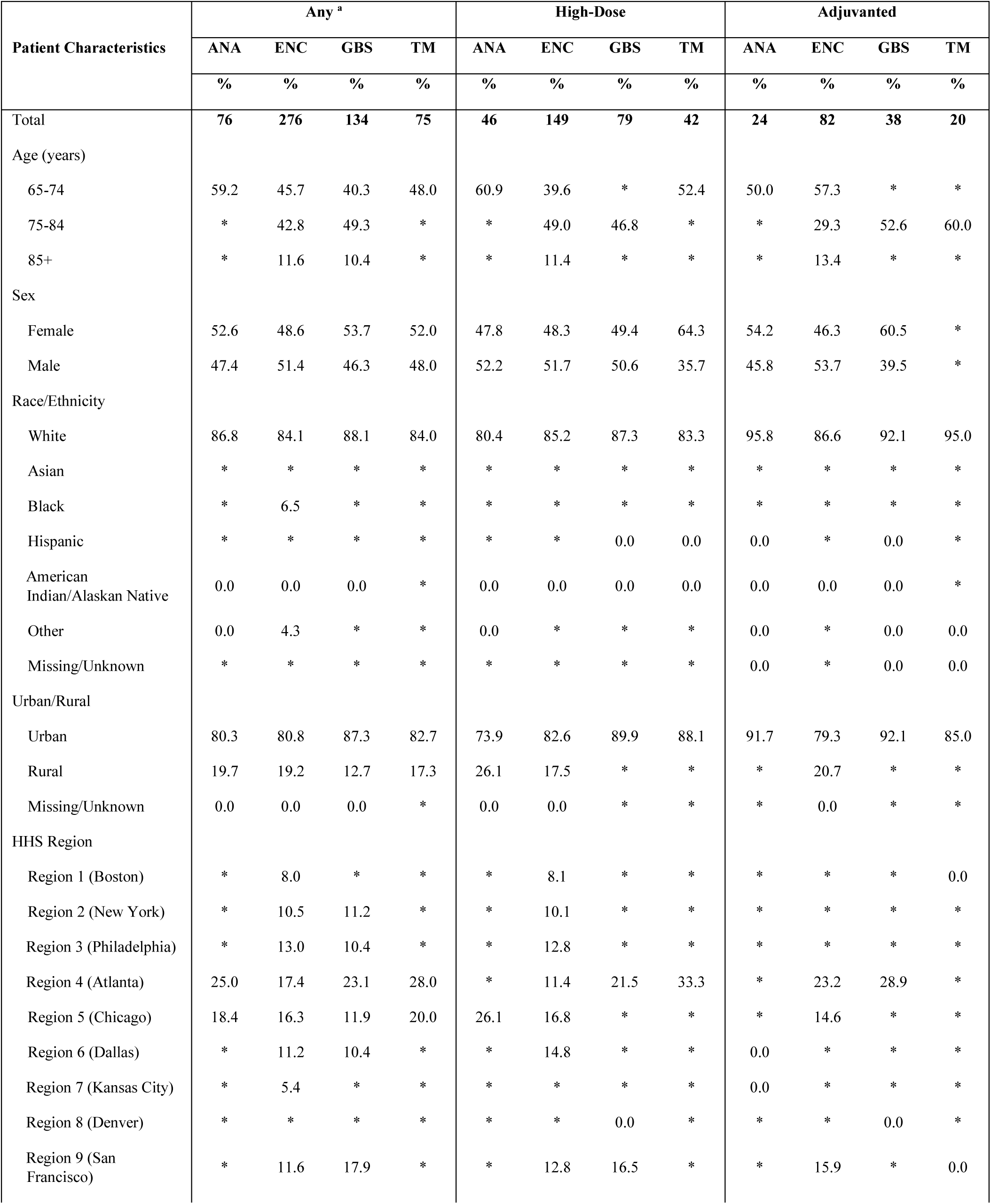
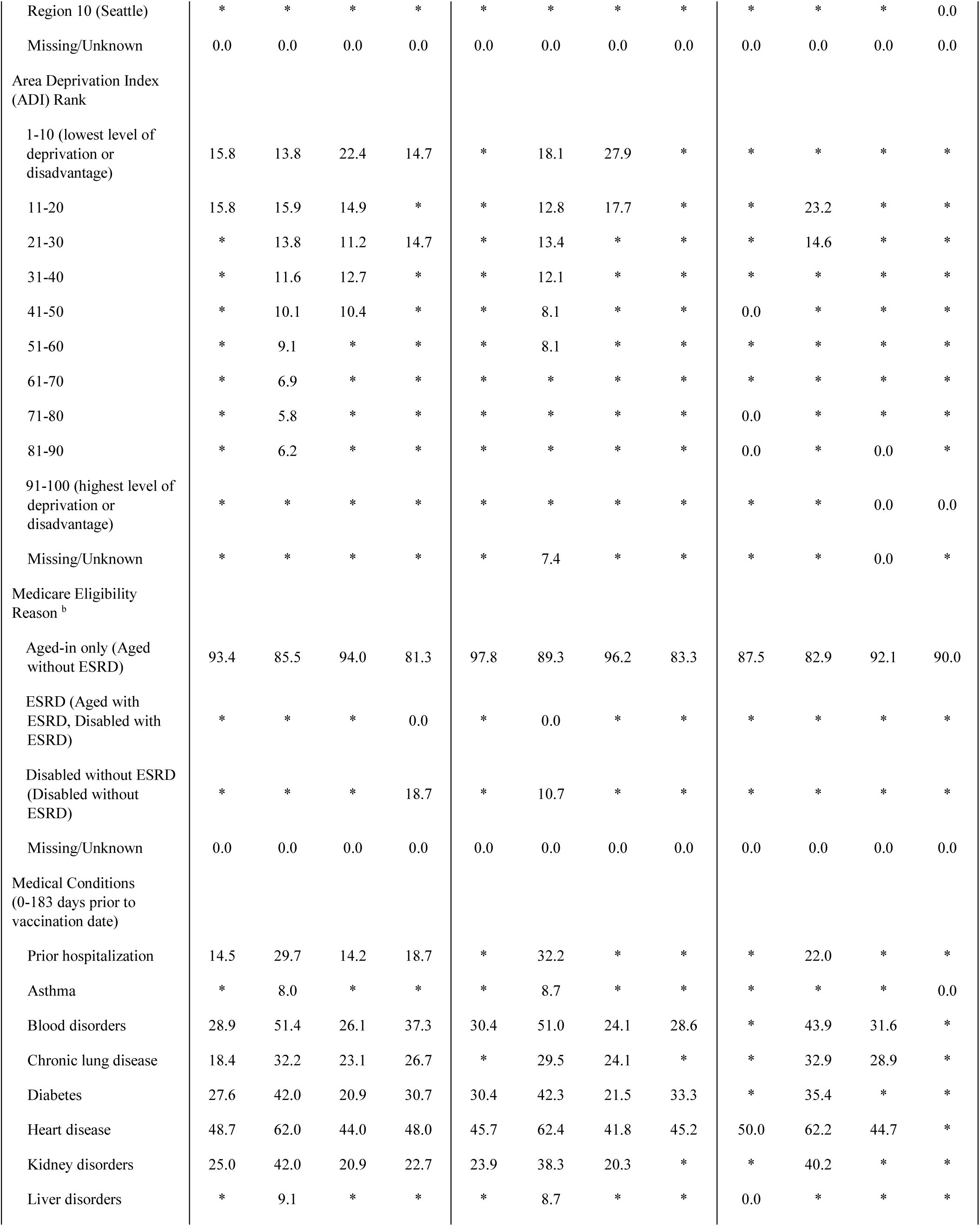
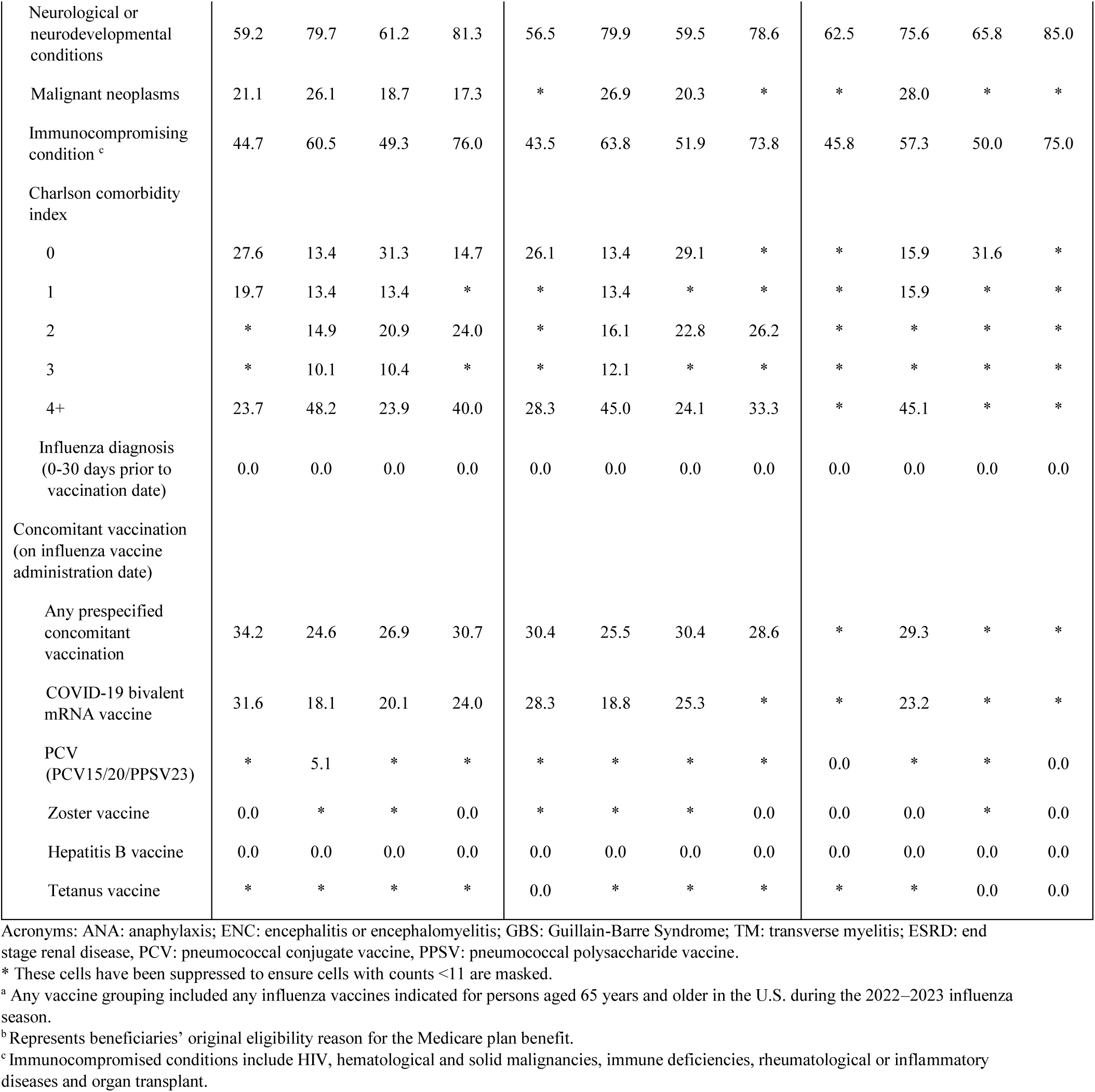
Description of the case population with each adverse event stratified by vaccine group.

### 3.2 Primary and Secondary Analyses

#### Anaphylaxis

In the seasonality and PPV-adjusted analysis, there was a statistically non-significant elevation in the rate of anaphylaxis following influenza vaccination for those receiving any (IRR: 2.40, 95% CI: 0.96–6.03), high-dose (IRR: 2.31, 95% CI: 0.67–7.91), or adjuvanted (IRR: 3.28, 95% CI: 0.71–15.08) influenza vaccines (Figures 1, Supplementary Table 5). However, in the seasonality-adjusted analysis, a statistically significant elevation in the rate was observed for those receiving any (IRR: 2.30, 95% CI: 1.24–4.27), high-dose (IRR: 2.24, 95% CI: 1.00–5.01) or adjuvanted (IRR: 3.17, 95% CI: 1.18–8.48) influenza vaccines (Supplementary Table 3). In the PPV-adjusted analysis, there was a statistically significant elevation in the rate for those receiving any influenza vaccine (IRR: 2.55, 95% CI: 1.02–6.40) (Supplementary Table 4). Anaphylaxis cases attributable to vaccination were rare (Supplementary Table 3–5).

**Figure 1.**
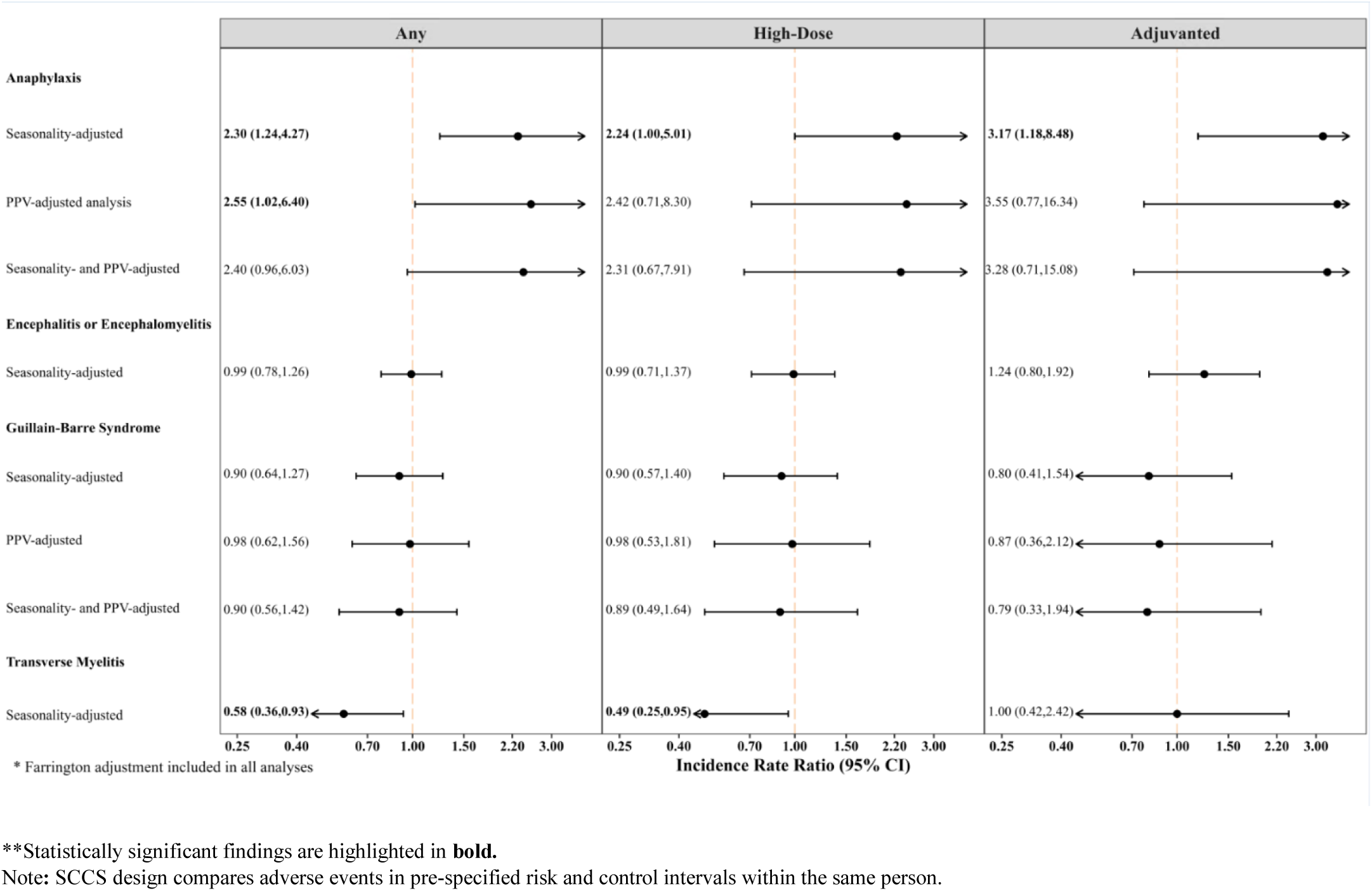
**Comparison of incidence rate ratios from primary self-controlled case series analyses across various bias-adjusted analyses, by high-dose and adjuvanted vaccine group**

In secondary analyses, seasonality and PPV-adjusted analyses found a statistically non-significant elevation in the anaphylaxis rate among those with any same-day concomitant vaccination (IRR: 2.54, 95% CI: 0.49–13.05) and without any same-day concomitant vaccination (IRR: 1.64, 95% CI: 0.38–7.05), for those receiving any influenza vaccination (Supplementary Table 8). Anaphylaxis cases attributable to influenza vaccination for both population subgroups with and without concomitant vaccination were also rare (Supplementary Tables 6–8).

#### Encephalitis/Encephalomyelitis

There was no association between the rate of encephalitis/encephalomyelitis among those receiving any (IRR: 0.99, 95% CI: 0.78–1.26), high-dose (IRR: 0.99, 95% CI: 0.71–1.37), or adjuvanted (IRR: 1.24, 95% CI: 0.80–1.92) influenza vaccines in the seasonality-adjusted analysis (Figure 1, Supplementary Table 3). Since a PPV estimate was not available for this outcome, PPV-adjusted analyses were not performed.

#### GBS

In the seasonality and PPV-adjusted analysis, there was no increase in the GBS rate following influenza vaccination for those receiving any (IRR: 0.90, 95% CI: 0.56–1.42), high-dose (IRR: 0.89, 95% CI: 0.49–1.64) or adjuvanted (IRR: 0.79, 95% CI: 0.33–1.94) influenza vaccines (Figure 1, Supplementary Table 5).

#### Transverse Myelitis

A statistically significant protective association for transverse myelitis was observed for those receiving any (IRR: 0.58, 95% CI: 0.36–0.93) or high-dose (IRR: 0.49, 95% CI: 0.25–0.95) influenza vaccines in the seasonality-adjusted analyses, but not for those receiving adjuvanted influenza vaccines (IRR: 1.00, 95% CI: 0.42–2.42) (Figure 1, Supplementary Table 3). A PPV estimate was not available for transverse myelitis; consequently, PPV-adjusted analyses were not performed.

In secondary analyses, a statistically non-significant protective association was found among those with concomitant vaccination (IRR: 0.64, 95% CI: 0.25–1.62) and without concomitant vaccination (IRR: 0.58, 95% CI: 0.31–1.09) for those receiving any influenza vaccination in seasonality-adjusted results (Supplementary Table 6).

However, in the post-hoc patient claims profile analysis, we found 76% of the 75 identified cases were unlikely incident idiopathic transverse myelitis cases based on the claims profile review (Table 3).

**Table 3.**
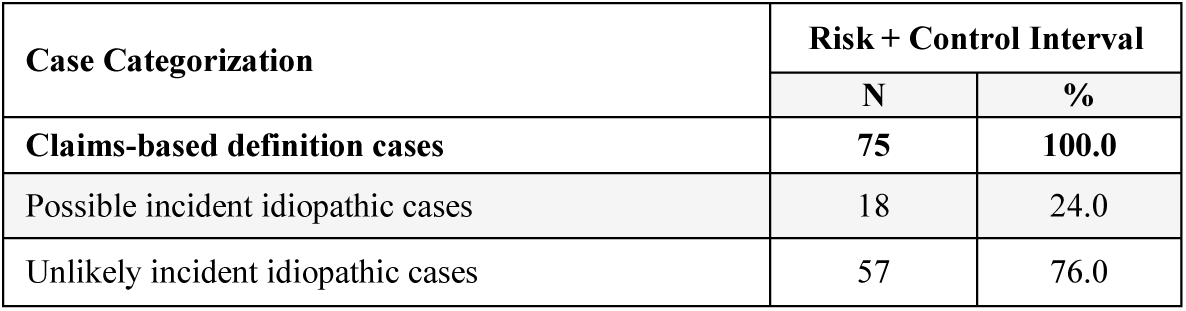
Case categorizations from transverse myelitis claims-based patient profile analysis.

**Table 4.** Incidence rate ratios from primary and secondary self-controlled case series analyses across various bias-adjustments, by vaccine group.

| Adverse Event | Bias-Adjustment | Analysis Type | Presence of Concomitant Vaccination <sup>a</sup> | Incidence Rate Ratio (95% CI) |
| --- | --- | --- | --- | --- |
| Anaphylaxis | Seasonality | Primary | NA | 2.30 (1.24, 4.27) |
|  |  | Secondary | Yes | 2.45 (0.84, 7.17) |
|  |  | Secondary | No | 1.62 (0.64, 4.11) |
|  | PPV | Primary | NA | 2.55 (1.02, 6.40) |
|  |  | Secondary | Yes | 2.69 (0.52, 13.87) |
|  |  | Secondary | No | 1.74 (0.40, 7.52) |
|  | Seasonality and PPV | Primary | NA | 2.40 (0.96, 6.03) |
|  |  | Secondary | Yes | 2.54 (0.49, 13.05) |
|  |  | Secondary | No | 1.64 (0.38, 7.05) |
| Transverse Myelitis | Seasonality | Primary | NA | 0.58 (0.36, 0.93) |
|  |  | Secondary | Yes | 0.64 (0.25, 1.62) |
|  |  | Secondary | No | 0.58 (0.31, 1.09) |

Acronyms: PPV: Positive predictive value; CI: confidence interval.
Note: Secondary analysis was only performed for adverse events with a statistically significant finding from the primary analysis. SCCS design compares adverse events in pre-specified risk and control intervals within the same person.
<sup>a</sup> Same-day concomitant vaccines include: COVID-19, PCV/PPSV, Zoster, Hepatitis B, and Tetanus.

## 4. Discussion

Our study of influenza vaccine safety in U.S. recipients aged 65 years and older found an elevated rate of anaphylaxis following receipt of any, high-dose, or adjuvanted influenza vaccines in a seasonality-adjusted analysis, which was consistent, but no longer statistically significant with adjustments for seasonality and outcome misclassification. No statistically significant elevation in rates was observed for encephalitis/encephalomyelitis, GBS, or transverse myelitis.

Case reports and surveillance studies of influenza vaccines from previous seasons have reported an increase in anaphylaxis risk^(16, 31, 32)^ while others have shown no association.^(7, 8)^ In our study, we observed an elevated rate of anaphylaxis following the 2022–2023 seasonal influenza vaccines recommended for those 65 years and older. However, the association was no longer statistically significant when analyses were adjusted for both seasonality and outcome misclassification. Some evidence has similarly shown a rare but elevated anaphylaxis risk associated with influenza vaccines from previous seasons.^(16, 31-33)^

The effect of concomitant vaccination on the association between influenza vaccines and anaphylaxis is still unclear. A non-statistically significant elevation in the rate of anaphylaxis was observed in population subgroups with and without any concomitant vaccinations, although the rate was attenuated in the population without any concomitant vaccination. Our main objective was not to study concomitant vaccination; therefore, we cannot distinguish the potential independent contribution of influenza vaccines, the bivalent COVID-19 mRNA vaccines, which were the most prevalent concomitant vaccines, or same-day coadministration of both vaccines on the possible increase in anaphylaxis incidence rates.

A protective effect for transverse myelitis was found following any or high-dose influenza vaccines. Several case reports have cited transverse myelitis cases following influenza vaccines.^(10, 13)^ However, no large-scale epidemiological studies, to our knowledge, have shown any association between transverse myelitis and influenza vaccination. While our study found a protective association for transverse myelitis following influenza vaccination, this result cannot be clinically or biologically justified. This result may have been due to bias from outcome misclassification.^(34)^ Our claims-based patient profile analysis found that over three fourths of transverse myelitis cases identified in the study were unlikely to be true incident idiopathic transverse myelitis cases. Instead, these cases tended to be secondary diagnoses unrelated to vaccination. We speculate that this was largely related to the International Classification of Diseases-10 diagnosis code used to identify transverse myelitis which is not specific to idiopathic transverse myelitis, thus likely leading to misclassification of unrelated cases as transverse myelitis cases in our study.

Our study used a self-controlled design which compared event incidence rates within rather than between individuals, and thus inherently adjusted for time-invariant confounders. Using PPVs from MRR for two of the four study outcomes helped to reduce bias from outcome misclassification on rate estimates. Similarly, our study included a large representative population of influenza vaccine recipients 65 years and older in the U.S. increasing the generalizability of the results to this population. The stratified analyses by concomitant vaccination status investigated the effect of concomitant vaccination on incidence rates of AEs. However, results were inconclusive, because of the limited sample size.

There are a few limitations to the study. Administrative claims databases were used to classify AEs; bias from outcome misclassification may therefore be present, particularly for encephalitis/encephalomyelitis and transverse myelitis, which did not undergo prior MRR. Prior influenza infection could have also biased rate estimates since influenza infection has been associated with certain study AEs.^(35-37)^ While we observed no prior influenza infections in the 30 days prior to vaccination among our cases, this may have been due to the limited observability of influenza diagnoses in claims based on the failure of affected persons to seek care and claims billing practices. In addition, it can be difficult to determine the precise timing when the rates of AEs post-vaccination return to baseline. Thus, there could be misspecification of the risk window that may result in residual bias.

This study was conducted by FDA in collaboration with CMS to evaluate the safety of the 2022–2023 seasonal influenza vaccines among U.S. adults 65 years and older. The study showed an elevation in anaphylaxis incidence rates associated with the influenza vaccine that could potentially be influenced by same-day concomitant vaccination. No statistically significant elevation in incidence rates of encephalitis /encephalomyelitis, GBS and transverse myelitis was associated with influenza vaccines. The FDA believes the benefits of seasonal influenza vaccination continue to outweigh the risks.

## Supporting information

Influenza Vaccine Safety Manuscript_Supplementary_Materials

## Data Availability

All data described in the present study is not available to protect patient confidentiality.

## Acknowledgements

We would like to thank Shwetha Krishnakumar, Jingjing An, Nimesh Shah, Xinxin (Kira) Lin, Meng Chen, and Zhiruo (Cassie) Wan for providing statistical programming and writing support.

## References

1. Centers for Disease Control and Prevention. Seasonal Flu Vaccines 2023 [Available from: https://www.cdc.gov/flu/prevent/flushot.htm#:~:text=For%20the%202022%2D2023%20flu%20season%2C%20there%20are%20three%20flu,Fluad%20Quadrivalent%20adjuvanted%20flu%20vaccine.

2. Langer J, Welch VL, Moran MM, Cane A, Lopez SMC, Srivastava A, et al. High Clinical Burden of Influenza Disease in Adults Aged ≥ 65 Years: Can We Do Better? A Systematic Literature Review. Advances in therapy. 2023;40(4):1601–27.

3. Centers for Disease Control and Prevention. Weekly Flu Vaccination Dashboard [cited 2023. Available from: https://www.cdc.gov/flu/fluvaxview/dashboard/vaccination-dashboard.html.

4. Centers for Disease Control and Prevention. What Vaccines are Recommended for You [cited 2023. Available from: https://www.cdc.gov/vaccines/adults/rec-vac/index.html.

5. Ghaderi S, Størdal K, Gunnes N, Bakken IJ, Magnus P, Håberg SE. Encephalitis after influenza and vaccination: a nationwide population-based registry study from Norway. International Journal of Epidemiology. 2017;46(5):1618–26.

6. Institute for Vaccine Safety JHBoSoPH. Do Vaccines Cause Transverse Myelitis? [cited 2023. Available from: https://www.vaccinesafety.edu/do-vaccines-cause-transverse-myelitis/.

7. Kawai AT, Li L, Kulldorff M, Vellozzi C, Weintraub E, Baxter R, et al. Absence of associations between influenza vaccines and increased risks of seizures, Guillain–Barré syndrome, encephalitis, or anaphylaxis in the 2012–2013 season. Pharmacoepidemiology & Drug Safety. 2014;23(5):548–53.

8. McCarthy NL, Gee J, Lin ND, Thyagarajan V, Pan Y, Su S, et al. Evaluating the safety of influenza vaccine using a claims-based health system. Vaccine. 2013;31(50):5975–82.

9. Yen C-C, Wei K-C, Wang W-H, Huang Y-T, Chang Y-C. Risk of Guillain-Barré Syndrome Among Older Adults Receiving Influenza Vaccine in Taiwan. JAMA Network Open. 2022;5(9):e2232571-e.

10. Akkad W, Salem B, Freeman JW, Huntington MK. Longitudinally Extensive Transverse Myelitis Following Vaccination With Nasal Attenuated Novel Influenza A(H1N1) Vaccine. Archives of Neurology. 2010;67(8):1018–20.

11. Bakshi R, Mazziotta JC. Acute transverse myelitis after influenza vaccination: magnetic resonance imaging findings. Journal of neuroimaging : official journal of the American Society of Neuroimaging. 1996;6(4):248–50.

12. Fujimori M, Nakamura M. Association between seasonal influenza vaccines and the increased risk of acute disseminated encephalomyelitis, estimated using the Vaccine Adverse Event Reporting System. Die Pharmazie. 2022;77(7):262–9.

13. Gui L, Chen K, Zhang Y. Acute transverse myelitis following vaccination against H1N1 influenza: a case report. International journal of clinical and experimental pathology. 2011;4(3):312–4.

14. Levison LS, Thomsen RW, Andersen H. Guillain-Barré syndrome following influenza vaccination: A 15-year nationwide population-based case-control study. European journal of neurology. 2022;29(11):3389–94.

15. Machicado JD, Bhagya-Rao B, Davogustto G, McKelvy BJ. Acute disseminated encephalomyelitis following seasonal influenza vaccination in an elderly patient. Clinical and vaccine immunology : CVI. 2013;20(9):1485–6.

16. Rouleau I, De Serres G, Drolet JP, Skowronski DM, Ouakki M, Toth E, et al. Increased risk of anaphylaxis following administration of 2009 AS03-adjuvanted monovalent pandemic A/H1N1 (H1N1pdm09) vaccine. Vaccine. 2013;31(50):5989–96.

17. Petersen I, Douglas I, Whitaker H. Self controlled case series methods: an alternative to standard epidemiological study designs. BMJ. 2016;354:i4515.

18. Daley MF, Clarke CL, Glanz JM, Xu S, Hambidge SJ, Donahue JG, et al. The safety of live attenuated influenza vaccine in children and adolescents 2 through 17 years of age: A Vaccine Safety Datalink study. 2018;27(1):59–68.

19. Yih WK, Kulldorff M, Sandhu SK, Zichittella L, Maro JC, Cole DV, et al. Prospective influenza vaccine safety surveillance using fresh data in the Sentinel System. 2016;25(5):481–92.

20. Food and Drug Administration. Master Protocol: Assessment of the Risk of Adverse Events Following Influenza Vaccinations Approved during the 2022-2023 Influenza Season 2023 [cited 2023. Available from: https://bestinitiative.org/wp-content/uploads/2023/06/BEST_2022-2023_Influenza-AE_Protocol_2023.pdf.

21. Baxter R, Lewis E, Goddard K, Fireman B, Bakshi N, DeStefano F, et al. Acute Demyelinating Events Following Vaccines: A Case-Centered Analysis. Clinical Infectious Diseases. 2016;63(11):1456–62.

22. Goud R, Lufkin B, Duffy J, Whitaker B, Wong HL, Liao J, et al. Risk of Guillain-Barré Syndrome Following Recombinant Zoster Vaccine in Medicare Beneficiaries. JAMA Intern Med. 2021;181(12):1623–30.

23. Shimabukuro TT, Cole M, Su JR. Reports of Anaphylaxis After Receipt of mRNA COVID-19 Vaccines in the US—December 14, 2020-January 18, 2021. JAMA. 2021;325(11):1101–2.

24. Souayah N, Nasar A, Suri MFK, Qureshi AI. Guillain-Barré Syndrome after Vaccination in United States: Data From the Centers for Disease Control and Prevention/Food and Drug Administration Vaccine Adverse Event Reporting System (1990-2005). Journal of Clinical Neuromuscular Disease. 2009;11(1).

25. Farrington CP, Whitaker HJ, Hocine MN. Case series analysis for censored, perturbed, or curtailed post-event exposures. Biostatistics (Oxford, England). 2009;10(1):3–16.

26. Hamedani AG, Thibault D, Willis AW. Seasonal Variation in Neurologic Hospitalizations in the United States. Annals of neurology. 2023;93(4):743–51.

27. Mulla ZD, Simon MR. Anaphylaxis in Olmsted County: Seasonal pattern and suggestions for epidemiologic analysis. Journal of Allergy and Clinical Immunology. 2009;123(5):1194.

28. Goud R, Thompson D, Welsh K, Lu M, Loc J, Lindaas A, et al. ICD-10 anaphylaxis algorithm and the estimate of vaccine-attributable anaphylaxis incidence in Medicare. Vaccine. 2021;39(38):5368–75.

29. Arya DP, Said MA, Izurieta HS, Perez-Vilar S, Zinderman C, Wernecke M, et al. Surveillance for Guillain-Barré syndrome after 2015–2016 and 2016–2017 influenza vaccination of Medicare beneficiaries. Vaccine. 2019;37(43):6543–9.

30. Rubin DB, Schenker N. Multiple Imputation for Interval Estimation From Simple Random Samples With Ignorable Nonresponse. Journal of the American Statistical Association. 1986;81(394):366–74.

31. Kim MJ, Shim DH, Cha HR, Kim CB, Kim SY, Park JH, et al. Delayed-Onset Anaphylaxis Caused by IgE Response to Influenza Vaccination. Allergy, asthma & immunology research. 2020;12(2):359–63.

32. McNeil MM, Weintraub ES, Duffy J, Sukumaran L, Jacobsen SJ, Klein NP, et al. Risk of anaphylaxis after vaccination in children and adults. Journal of Allergy and Clinical Immunology. 2016;137(3):868–78.

33. McNeil MM. Vaccine-Associated Anaphylaxis. Current treatment options in allergy. 2019;6(3):297–308.

34. Williams SE, Carnahan R, Krishnaswami S, McPheeters ML. A systematic review of validated methods for identifying transverse myelitis using administrative or claims data. Vaccine. 2013;31 Suppl 10:K83–7.

35. Márquez S, Vilá LM. Extensive Longitudinal Transverse Myelitis after Influenza A Virus Infection in a Patient with Systemic Lupus Erythematosus. Case reports in rheumatology. 2022;2022:9506733.

36. Sellers SA, Hagan RS, Hayden FG, Fischer WA, 2nd. The hidden burden of influenza: A review of the extra-pulmonary complications of influenza infection. Influenza and other respiratory viruses. 2017;11(5):372–93.

37. Sivadon-Tardy V, Orlikowski D, Porcher R, Sharshar T, Durand MC, Enouf V, et al. Guillain-Barré syndrome and influenza virus infection. Clinical infectious diseases : an official publication of the Infectious Diseases Society of America. 2009;48(1):48–56.

