## Supplementary material for "Assessment of Potential Adverse Events Following the 2022–2023 Seasonal Influenza Vaccines Among U.S. Adults Aged 65 Years and Older": Influenza Vaccine Safety Manuscript_Supplementary_Materials

**Supplementary Table 1. Description of Claims-Based Patient Profile Analysis Method and Criteria Used for Classification of “Possible” and “Unlikely” Incident Idiopathic Cases**

**Claims Patient Profile Analysis Method**

The patient profile analysis involved clinician review of longitudinal claims profiles for all 75 transverse myelitis cases in inpatient and outpatient facility settings, and physician services settings where applicable. For each case, claims profiles were created using claims from inpatient, outpatient facility and physician services claims settings, and were based on a lookback period of 6 months prior to the incident outcome diagnosis and a lookforward of at least 2 months following the incident outcome diagnosis. Each profile included medical diagnosis and procedural codes (ICD-10, CPT/HCPCS), and their associated description, provider type, care setting, and billing date. Billing dates were displayed relative to the index transverse myelitis diagnosis. Our clinician team including input from a neurologist developed predefined criteria that were used to determine if transverse myelitis cases identified from the claims-based definition were “possible” or “unlikely” incident idiopathic cases (Table A1).^(1-3)^ Incident idiopathic cases are a subset of transverse myelitis cases that we expect to be possibly attributable to vaccination rather than potential unrelated causes. Reviewing clinicians were blinded to the intervals in which these cases were identified in the study (i.e., risk versus control intervals).

**Criteria to Identify Possible Incident Idiopathic Transverse Myelitis Cases**

**(i) Do not have any of the exclusion criteria below**

(a) Presence of ICD-10 codes for conditions which may be causes of transverse myelitis e.g., multiple sclerosis, neuromyelitis optica disorder, infections, nutritional conditions, autoimmune conditions, radiation, vascular insufficiency of the spinal cord, and trauma to the spine.

**(ii) Have at least 3 of the inclusion criteria below**

(a) No prior diagnosis of transverse myelitis in the 6 months lookback in any care settings.
(b) Persistent diagnosis of transverse myelitis in the claims profile after the incident case, in all care settings (at least 2 diagnosis required in each setting).
(c) Procedure codes for magnetic resonance imaging/cerebrospinal fluid examination/ plasmapheresis/ Foley catheterization, in claims profile within 15 days of the incident transverse myelitis diagnosis.
(d) Consultation by a neurologist in both inpatient and outpatient facility settings.
(e) Inpatient hospitalization during the diagnosis of incident idiopathic transverse myelitis.

**Supplementary Table 2.** **Population exclusion table for AE-specific cohorts**

| Eligibility Criteria | Anaphylaxis | | Encephalitis or Encephalomyelitis | | Guillain-Barré Syndrome | | Transverse Myelitis | |
| --- | --- | --- | --- | --- | --- | --- | --- | --- |
|  | **No.** | **%** | **No.** | **%** | **No.** | **%** | **No.** | **%** |
| Received a dose of influenza vaccine during study period prior to vaccination cutoff ^a^ | 12,683,255 | 100.0 | 12,683,255 | 100.0 | 12,683,255 | 100.0 | 12,683,255 | 100.0 |
| Enrolled in Part A and Part B at the time of vaccination | 12,375,116 | 97.6 | 12,375,116 | 97.6 | 12,375,116 | 97.6 | 12,375,116 | 97.6 |
| At least 65 yrs. old at the time of vaccination | 11,661,367 | 91.9 | 11,661,367 | 91.9 | 11,661,367 | 91.9 | 11,661,367 | 91.9 |
| Did not have multiple influenza vaccines on the same day or within 3 days of each | 11,650,128 | 91.9 | 11,650,128 | 91.9 | 11,650,128 | 91.9 | 11,650,128 | 91.9 |
| Continuously enrolled in Medicare Parts A/B FFS 365 days prior to vaccination | 10,976,879 | 86.5 | 10,976,879 | 86.5 | 10,976,879 | 86.5 | 10,976,879 | 86.5 |
| Continuously enrolled in Medicare Parts A/B FFS 365 days from start of observation period until end of observation period/death/disenrollment/end of study period/subsequent influenza vaccination | 10,976,514 | 86.5 | 10,976,091 | 86.5 | 10,976,091 | 86.5 | 10,976,091 | 86.5 |
| Experienced the AE during the observation period | 76 | 0.0 | 313 | 0.0 | 143 | 0.0 | 99 | 0.0 |
| Did not receive an AE diagnosis during the clean interval | 76 | 0.0 | 294 | 0.0 | 140 | 0.0 | 76 | 0.0 |
| Had follow-up time in both risk and control intervals | 76 | 0.0 | 276 | 0.0 | 134 | 0.0 | 75 | 0.0 |
| Case Population | **76** | | **276** | | **134** | | **75** | |

Acronyms: AE: Adverse event; FFS: Fee-for-service (Medicare).
^a^ Beneficiaries with an influenza vaccine administration between 8/1/2022 and 11/12/2022.

**Supplementary Table 3. Incidence rate ratio and attributable risk estimates adjusted for seasonality, stratified by vaccine group**

| Adverse Event | Influenza Vaccine Group | Risk Interval ^a^ | | Control Interval | | IRR (95% CI) | No. of  Vaccinated  Individuals | Person-Years | AR ^b^ (95% CI) |
| --- | --- | --- | --- | --- | --- | --- | --- | --- | --- |
|  |  | **No. Cases** | **Person-Days** | **No. Cases** | **Person-Days** |  |  |  |  |
| Anaphylaxis | Any | 12 | 152 | 64 | 1,140 | 2.30 (1.24,4.27) | 10,976,514 | 60,143 | 0.06 (0.00,0.12) |
|  | High-Dose | * | 92 | 39 | 690 | 2.24 (1.00, 5.01) | 6,548,717 | 35,882 | 0.06 (-0.02,0.14) |
|  | Adjuvanted | * | 48 | 19 | 360 | 3.17 (1.18,8.48) | 3,406,199 | 18,664 | 0.10 (-0.02,0.22) |
| Encephalitis or Encephalomyelitis | Any | 137 | 12,128 | 157 | 12,248 | 0.99 (0.78,1.26) | 10,976,091 | 1,259,278 | -0.01 (-0.31,0.29) |
|  | High-Dose | 72 | 6,507 | 85 | 6,629 | 0.99 (0.71,1.37) | 6,548,563 | 751,624 | -0.01 (-0.37,0.34) |
|  | Adjuvanted | 46 | 3,634 | 42 | 3,655 | 1.24 (0.80,1.92) | 3,406,105 | 390,913 | 0.26 (-0.26,0.78) |
| Guillain-Barré Syndrome | Any | 66 | 5,784 | 74 | 6,271 | 0.90 (0.64,1.27) | 10,976,091 | 1,259,278 | -0.06 (-0.28,0.15) |
|  | High-Dose | 40 | 3,445 | 44 | 3,726 | 0.90 (0.57,1.40) | 6,548,563 | 751,624 | -0.07 (-0.36,0.22) |
|  | Adjuvanted | 17 | 1,625 | 22 | 1,780 | 0.80 (0.41,1.54) | 3,406,105 | 390,913 | -0.13 (-0.49,0.24) |
| Transverse Myelitis | Any | 27 | 3,150 | 48 | 3,536 | 0.58 (0.36,0.93) | 10,976,091 | 1,259,278 | -0.18 (-0.32,-0.03) |
|  | High-Dose | 12 | 1,764 | 30 | 1,980 | 0.49 (0.25,0.95) | 6,548,563 | 751,624 | -0.19 (-0.38,-0.01) |
|  | Adjuvanted | * | 840 | * | 937 | 1.00 (0.42,2.42) | 3,406,105 | 390,913 | 0.00 (-0.25,0.25) |

Acronyms: CI: confidence interval, IRR: incidence rate ratio; AR: attributable risk.
Note: SCCS design compares adverse events in pre-specified risk and control intervals within the same person. High potential rate of outcome misclassification was identified for transverse myelitis outcome; statistically significant finding is thus not conclusive.

^a^ Incident cases occurring in the risk window regardless of if accrued control time. Cases with multiple influenza vaccines in the risk interval were excluded.

^b^ per 100,000 vaccinations.
* These cells have been suppressed to ensure cells with counts <11 are masked.

**Supplementary Table 4. Incidence rate ratio and attributable risk estimates adjusted for adverse event-specific PPV, stratified by vaccine group**

| Adverse Event | Influenza Vaccine Group | Risk Interval | | Control Interval | | IRR (95% CI) | No. of Vaccinated Individuals | Person-Years | AR ^a^ (95% CI) |
| --- | --- | --- | --- | --- | --- | --- | --- | --- | --- |
|  |  | **No. Cases** | **Person-Days** | **No. Cases** | **Person-Days** |  |  |  |  |
| Anaphylaxis | Any | 7.88 | 101 | 42.48 | 755 | 2.55 (1.02,6.40) | 10,976,514 | 60,143 | 0.04 (-0.02,0.10) |
|  | High-Dose | 4.61 | 61 | 25.82 | 456 | 2.42 (0.70,8.30) | 6,548,717 | 35,882 | 0.04(-0.04,0.12) |
|  | Adjuvanted | 3.34 | 32 | 12.55 | 238 | 3.55 (0.77,16.34) | 3,406,199 | 18,664 | 0.07 (-0.05,0.19) |
| Guillain-Barré Syndrome | Any | 46.75 | 4,109 | 52.71 | 4,456 | 0.98 (0.62,1.56) | 10,976,091 | 1,259,278 | -0.01 (-0.21,0.20) |
|  | High-Dose | 28.46 | 2,452 | 31.32 | 2,652 | 0.98 (0.53,1.81) | 6,548,563 | 751,624 | -0.01 (-0.28,0.27) |
|  | Adjuvanted | 12.06 | 1,155 | 15.66 | 1,265 | 0.87 (0.36,2.12) | 3,406,105 | 390,913 | -0.05 (-0.40,0.30) |

Acronyms: PPV: positive predictive value; CI: confidence interval, IRR: incidence rate ratio; AR: attributable risk.

Note: SCCS design compares adverse events in pre-specified risk and control intervals within the same person.

^a^ per 100,000 vaccinations.

**Supplementary Table 5. Incidence rate ratio and attributable risk estimates adjusted for seasonality and PPV, stratified by vaccine group**

| Adverse Event | Influenza Vaccine Group | Risk Interval | | Control Interval | | IRR (95% CI) | No. of Vaccinated Individuals | Person-Years | AR ^a^ (95% CI) |
| --- | --- | --- | --- | --- | --- | --- | --- | --- | --- |
|  |  | **No. Cases** | **Person-Days** | **No. Cases** | **Person-Days** |  |  |  |  |
| Anaphylaxis | Any | 7.88 | 101 | 42.48 | 755 | 2.40 (0.96, 6.03) | 10,976,514 | 60,143 | 0.04 (-0.02, 0.10) |
|  | High-Dose | 4.61 | 61 | 25.82 | 456 | 2.31 (0.67, 7.91) | 6,548,717 | 35,882 | 0.04 (-0.04, 0.12) |
|  | Adjuvanted | 3.34 | 32 | 12.55 | 238 | 3.28 (0.71, 15.08) | 3,406,199 | 18,664 | 0.07 (-0.05, 0.19) |
| Guillain-Barré Syndrome | Any | 46.75 | 4,109 | 52.71 | 4,456 | 0.90 (0.56, 1.42) | 10,976,091 | 1,259,278 | -0.05 (-0.27, 0.17) |
|  | High-Dose | 28.46 | 2,452 | 31.32 | 2,652 | 0.89 (0.49, 1.64) | 6,548,563 | 751,624 | -0.05 (-0.34, 0.24) |
|  | Adjuvanted | 12.06 | 1,155 | 15.66 | 1,265 | 0.79 (0.33, 1.94) | 3,406,105 | 390,913 | -0.09 (-0.46, 0.28) |

Acronyms: PPV: positive predictive value; CI: confidence interval, IRR: incidence rate ratio; AR: attributable risk.
Note: SCCS design compares adverse events in pre-specified risk and control intervals within the same person.

^a^ per 100,000 vaccinations.

**Supplementary Table 6**. **Incidence rate ratio and attributable risk estimates adjusted for seasonality for overall influenza vaccine group, stratified by concomitant vaccination status**

| Adverse Event | Concomitant Vaccination Status ^a^ | Risk Interval ^b^ | | Control Interval | | IRR (95% CI) | No. of Vaccinated Individuals | Person-Years | AR ^c^ (95% CI) |
| --- | --- | --- | --- | --- | --- | --- | --- | --- | --- |
|  |  | **No. Cases** | **Person-Days** | **No. Cases** | **Person-Days** |  |  |  |  |
| Anaphylaxis | With Concomitant Vaccinations | * | 48 | 20 | 360 | 2.45 (0.84, 7.17) | 2,210,186 | 12,110 | 0.11 (-0.06, 0.28) |
|  | No Concomitant Vaccinations | * | 86 | 38 | 645 | 1.62 (0.64, 4.11) | 6,690,312 | 36,658 | 0.03 (-0.04, 0.09) |
| Transverse Myelitis | With Concomitant Vaccinations | * | 798 | 12 | 889 | 0.64 (0.25,1.62) | 2,207,055 | 253,453 | -0.18 (-0.56, 0.20) |
|  | No Concomitant Vaccinations | 15 | 1,806 | 28 | 2,037 | 0.58 (0.31, 1.09) | 6,681,640 | 766,414 | -0.16 (-0.35, 0.02) |

Acronyms: CI: confidence interval, IRR: incidence rate ratio; AR: attributable risk.

Note: SCCS design compares adverse events in pre-specified risk and control intervals within the same person. High potential rate of outcome misclassification was identified for transverse myelitis outcome; statistically significant finding is thus not conclusive.
*These cells have been suppressed to ensure cells with counts <11 are masked.

^a^ Same-day concomitant vaccines include: COVID-19, PCV/PPSV, Zoster, Hepatitis B, and Tetanus.

^b^ Incident cases occurring in the risk window regardless of if accrued control time. Cases with multiple influenza vaccines in the risk interval were excluded.

^c^ per 100,000 vaccinations.

**Supplementary Table 7**. **Incidence rate ratio and attributable risk estimates adjusted for PPV for overall influenza vaccine group, stratified by concomitant vaccination status**

| Adverse Event | Concomitant Vaccination Status ^a^ | Risk Interval | | Control Interval | | IRR (95% CI) | No. of Vaccinated Individuals | Person-Years | AR ^b^ (95% CI) |
| --- | --- | --- | --- | --- | --- | --- | --- | --- | --- |
|  |  | **No. Cases** | **Person-Days** | **No. Cases** | **Person-Days** |  |  |  |  |
| Anaphylaxis | With Concomitant Vaccinations | 2.69 | 32 | 13.23 | 239 | 2.69 (0.52, 13.87) | 2,210,186 | 12,110 | 0.08 (-0.09, 0.25) |
|  | No Concomitant Vaccinations | 3.30 | 57 | 25.18 | 427 | 1.74 (0.40, 7.52) | 6,690,312 | 36,658 | 0.02 (-0.04, 0.09) |

Acronyms: PPV: positive predictive value; CI: confidence interval, IRR: incidence rate ratio; AR: attributable risk.
Note: SCCS design compares adverse events in pre-specified risk and control intervals within the same person.

^a^ Same-day concomitant vaccines include: COVID-19, PCV/PPSV, Zoster, Hepatitis B, and Tetanus vaccines.

^b^ per 100,000 vaccinations.

**Supplementary Table 8**. **Incidence rate ratio and attributable risk estimates adjusted for seasonality and PPV for overall influenza vaccine group, stratified by concomitant vaccination status**

| Adverse Event | Concomitant Vaccination Status ^a^ | Risk Interval | | Control Interval | | IRR (95% CI) | No. of Vaccinated Individuals | Person-Years | AR ^b^ (95% CI) |
| --- | --- | --- | --- | --- | --- | --- | --- | --- | --- |
|  |  | **No. Cases** | **Person-Days** | **No. Cases** | **Person-Days** |  |  |  |  |
| Anaphylaxis | With Concomitant Vaccinations | 2.69 | 32 | 13.23 | 239 | 2.54 (0.49, 13.05) | 2,210,186 | 12,110 | 0.07 (-0.10, 0.24) |
|  | No Concomitant Vaccinations | 3.30 | 57 | 25.18 | 427 | 1.64 (0.38, 7.05) | 6,690,312 | 36,658 | 0.02 (-0.05, 0.09) |

Acronyms: PPV: positive predictive value; CI: confidence interval, IRR: incidence rate ratio; AR: attributable risk.
Note: SCCS design compares adverse events in pre-specified risk and control intervals within the same person.

^a^ Same-day concomitant vaccines include: COVID-19, PCV/PPSV, Zoster, Hepatitis B, and Tetanus vaccines.

^b^ per 100,000 vaccinations.

**References**

1.	Cleveland Clinic. Transverse Myelitis [updated 2022. Available from: <https://my.clevelandclinic.org/health/diseases/8980-transverse-myelitis>.

2.	Transverse Myelitis Consortium Working Group. Proposed diagnostic criteria and nosology of acute transverse myelitis. Neurology. 2002;59(4):499-505.

3.	West TW, Hess C, Cree BA. Acute transverse myelitis: demyelinating, inflammatory, and infectious myelopathies. Seminars in neurology. 2012;32(2):97-113.
